# Association of single nucleotide polymorphisms in *XRCC1* and *ATM* with recurrence in patients with cervical cancer

**DOI:** 10.1101/2021.04.26.21256088

**Authors:** Soo Youn Cho, Kidong Kim, Sang Young Ryu, Moon-Hong Kim, Beob-Jong Kim

## Abstract

**Objective:** The primary objective of this study was to determine the association of the genotype of X-ray repair complementing defective repair in Chinese hamster cell 1 *(XRCC1)* 580C>T with recurrence-free survival (RFS) in patients with cervical cancer who underwent radical surgery followed by adjuvant concurrent chemoradiation (CRT). Secondary objectives were to assess 1) the association of single nucleotide polymorphisms (SNPs) in *XRCC1* and ataxia telangiectasia mutated *(ATM)* gene with RFS, overall survival (OS) and completion of CRT, 2) the association of haplotypes of *XRCC1* and *ATM* with RFS, OS and completion of CRT, 3) concordance rate between genotypes from cancer tissue and that from normal tissue and 4) the association of clinicopathologic variables with RFS and OS.

**Materials and Methods:** Clinicopathologic variables were extracted from medical records of 159 patients who underwent radical hysterectomy followed by adjuvant CRT in a cancer center between 1999 and 2004. After exclusion of six patients without paraffin blocks, 153 patients were included in this study. DNA was extracted from a formalin-fixed, paraffin embedded tissue block containing tumor tissue and another block containing uninvolved lymph node. Genotyping for *XRCC1* 580C>T (rs 1799782), *XRCC1* 1196A>G (rs 25487), *ATM* 3078-77C>T (rs 664677), *ATM* 5557G>A (rs 1801516) and *ATM* 6807+238G>C (rs609429) was performed using pyrosequencing. Haplotype estimation was performed using haplo.stats in R version 2.15.0. The association of genotype or haplotype of SNPs with RFS, OS and completion of CRT was evaluated via Cox regression analysis, chi-square or Fisher’s exact test. The concordance rate between genotype from cancer tissue and that from normal tissue was examined according to SNP. The association of clinicopathologic variables with RFS and OS was estimated via Cox regression analysis.

**Results:** None of SNPs was associated with RFS, OS and completion of CRT. Concordance rate between genotype from cancer tissue and that from normal tissue for *XRCC1* 580C>T, *XRCC1* 1196A>G, *ATM* 3078-77C>T, *ATM* 5557G>A and *ATM* 6807+238G>C were 72%, 88%, 73%, 100% and 54%, respectively. Age, stage, histologic type, LVSI and parametrial invasion were associated with RFS, and clinical tumor size, resection margin, lymph node involvement and completion of CRT were associated with OS.

**Conclusion:** Genotype of *XRCC1* 580C>T was not associated with RFS in patients with cervical cancer who underwent radical surgery followed by adjuvant CRT. Low concordance rate between cancer and germline genotype was identified in cervical cancer.

## Introduction

Cervical cancer is one of the common cancers in women. According to GLOBOCAN 2002, the age-standardized incidence of cervical cancer was 0.56 per 100,000 person-years and was the second most common cancer in women worldwide. ^1^ The treatment of cervical cancer differs according to the extent of disease. For early stage cervical cancer, radical surgery is usually performed, but for advanced cervical cancer, radiation with or without chemotherapy is the standard treatment. Among patients with early stage cervical cancer who initially received radical surgery, high- or intermediate-risk patients should undergo adjuvant treatment. As an adjuvant treatment, concurrent chemoradiation (CRT) is indicated for high-risk patients2 and radiation alone or CRT is performed for intermediate-risk patients. ^3,4^

However, a significant number of patients suffer recurrences despite adjuvant treatment. The literature shows that projected recurrence-free survival (RFS) of high-risk patients who had received radical surgery followed by adjuvant CRT was 80% ^2^ and recurrence-free rates at two years for intermediate-risk patients who had received radical surgery followed by adjuvant radiation was 88%. ^5^ Because the gross tumor is completely resected in most cases of early stage cervical cancer, the resistance of cancer cells to adjuvant treatment would be the major mechanism of recurrence.

Single nucleotide polymorphism (SNP) is a polymorphism in which the minor variant (allele) is present in at least 1% of a given population and account for most of the known genetic variation. ^6^ SNPs can affect protein function thorough various mechanisms and are associated with response to treatment and survival in patients with cancer. ^7^ I previously reported that the genotype of X-ray repair complementing defective repair in Chinese hamster cell 1 *(XRCC1)* 580C>T SNP was associated with response to platinum-based, neoadjuvant chemotherapy in cervical cancer. ^8^ Because adjuvant CRT after radical surgery consisted of platinum-based chemotherapy and pelvic radiation, I hypothesized that genotype of *XRCC1* 580C>T SNP could be associated with RFS in patients with cervical cancer who underwent radical surgery followed by adjuvant CRT.

The primary objective of the current study was to determine whether the genotype of *XRCC1* 580C>T is associated with RFS in patients with cervical cancer who underwent radical surgery followed by adjuvant CRT. Secondary objectives were to assess 1) the association of SNPs in *XRCC1* and ataxia telangiectasia mutated *(ATM)* gene with RFS, overall survival (OS) and completion of CRT, 2) the association of haplotypes of *XRCC1* and *ATM* with RFS, OS and completion of CRT, 3) concordance rate between genotype from cancer tissue and that from normal tissue and 4) the association of clinicopathologic variables with RFS and OS. Genotype determined by pyrosequencing using cancer tissue was intended to be used for all analysis. However, in case of significant discordance between genotype from cancer tissue and that from normal tissue, the association of genotype from normal tissue with RFS, OS and completion of CRT was planned to be examined.

## Materials and methods

### Design of study

The null hypothesis for sample size calculation was “the 2-year RFS of patients with CC genotype of *XRCC1* 580C>T is equal to that of patients with CT or TT genotype.” The ratio between CC vs. CT or TT genotype in my previous study was 52:48.^8^ Therefore, the ratio between CC vs. CT or TT genotype was assumed as 1:1. To estimate 2-year RFS according to genotypes, the RFS of 27 patients who were included in my previous study^8^ and received adjuvant CRT after radical surgery were estimated. The estimated 2-year RFS of patients with CC genotype was 85.9% and that of patients with CT or TT genotype was 63.6%. A difference in RFS of 22.3% (85.9% minus 63.6%) is considered a clinically important difference to detect. This difference require 63 patients in each genotype group to afford statistical power of at least 0.80 when keeping the probability of type 1 error at significance level, α=0.05. Considering the 10% failure rate to extract DNA from paraffin block or to determine genotypes, 70 patients in each genotype group is required.

Through searching the patient registry, I identified 159 patients who underwent radical hysterectomy followed by adjuvant CRT between 1999 and 2004. Because the number of patients identified (n=159) was similar to the statistically required number of patients (n=140), I decided to include all patients identified. Of 159 patients, six patients were excluded because both cancer and normal tissue blocks were unavailable. The remaining 153 patients were included in this study. Of 153 patients, 104 patients had both cancer and normal tissue blocks. Six patients had only normal tissue blocks and 43 patients had only cancer tissue blocks. After approval by the Institutional Review Board (K-1101-002-058), I abstracted clinicopathologic variables from medical records. Patients with parametrial invasion, positive resection margin or lymph node involvement were determined as high-risk, and those with intermediate-risk factors were determined as intermediate-risk based on the criteria used in a previous clinical trial. ^5^ RFS was defined as the length of time from the date of surgery to the date of recurrence. Likewise, OS was defined as the length of time from the date of surgery to the date of death from any cause.

### Selection of genes and SNPs

Because most adjuvant CRT comprised of platinum-based chemotherapy and radiation, I decided to choose one gene related to response to platinum-based chemotherapy and another gene related to the response to radiation. In my previous study, SNPs at *XRCC1* and gamma-glutamyl hydrolase *(GGH)* were associated with response to platinum-based chemotherapy. Because I thought that *XRCC1* is more consistently reported to be associated with response to platinum-based chemotherapy than *GGH* in literature, I selected XRCC1 for the current study.

A recent review summarized genes reported to be associated with response to radiation in various cancers. According to the review, SNPs in nine genes were associated with response to radiation: *ATM*, checkpoint kinase 1 *(CHK1)*, excision repair cross-complementing rodent repair deficiency complementation group 1 *(ERCC1), ERCC2, ERCC5*, Mdm2 p53 binding protein mouse homolog *(MDM2)*, tumor protein p53 *(TP53)*, xeroderma pigmentosum complementation group A *(XPA), XRCC1*.^7^ Because patients in the current study underwent CRT, I excluded genes *(ERCC5, XPA)* associated with response to radiation alone.^9^ Because the endpoint of the current study was RFS, I excluded a gene *(ERCC1)* which was not associated with RFS but with response rate.^10^ Genes *(ERCC2, MDM2, TP53)* associated with response to radiation only when they were combined with SNPs in other genes were excluded.^11,12^ *XRCC1* which is already selected for the current study was excluded. Between the remaining pair *ATM* and *CHK1*, I selected *ATM* because I thought that *ATM* is more consistently reported to be associated with response to radiation than *CHK1* in literature.

A gene function could be affected by multiple SNPs and would be better predicted by a combination of multiple SNPs than single SNP.^6^ Therefore, I aimed to select SNPs which could accurately determine haplotypes in >80% of cases. For *XRCC1*, based on a study including 229 Korean patients with lung cancer,^13^ 580C>T (rs 1799782), and 1196A>G (rs 25487) were chosen. For an *ATM* gene, there is no study investigating the frequency of each genotype of SNPs in Korean population. Therefore, I used the results of a study^14^ including 556 French women and selected 3078-77C>T (rs 664677), 5557G>A (rs 1801516) and 6807+238G>C (rs 609429).

### DNA extraction and SNP typing

In each patient, a formalin-fixed, paraffin embedded tissue block containing tumor tissue and another block containing uninvolved lymph node were chosen by a single pathologist (Cho SY). Three to five sections (10μm thick) were cut from each block and cells were scraped from the representative tumor portion or uninvolved lymph node. Scraped cells were stored at 4 °C until DNA extraction. QIAamp DNA Micro kit (Qiagen, Hilden, Germany) was used for DNA extraction according to the manufacturer’s instructions. The amount of DNA was measured by absorbance at 260 nm using an ultraviolet spectrometer, and the purity of the DNA was evaluated by measuring the 260/280 absorbance ratios.

Polymerase chain reaction (PCR) amplification of extracted DNA was performed with 50mM MgCl2 0.75μl, 25mM dNTP 0.2μl, 20pM forward and biotinylated reverse primers 0.5μl each, 5U/μl Taq polymerase 0.15μl, and 2.5μl DNA in a total volume of 25μl. The used primers for each SNP were summarized in Table 1. PCR conditions were 95°C for 10 min followed by 40 cycles (95°C for 30 seconds, 55°C for 30 seconds, 72°C for 30 seconds) and a final elongation step at 72°C for 5 minutes.

**Table 1.**
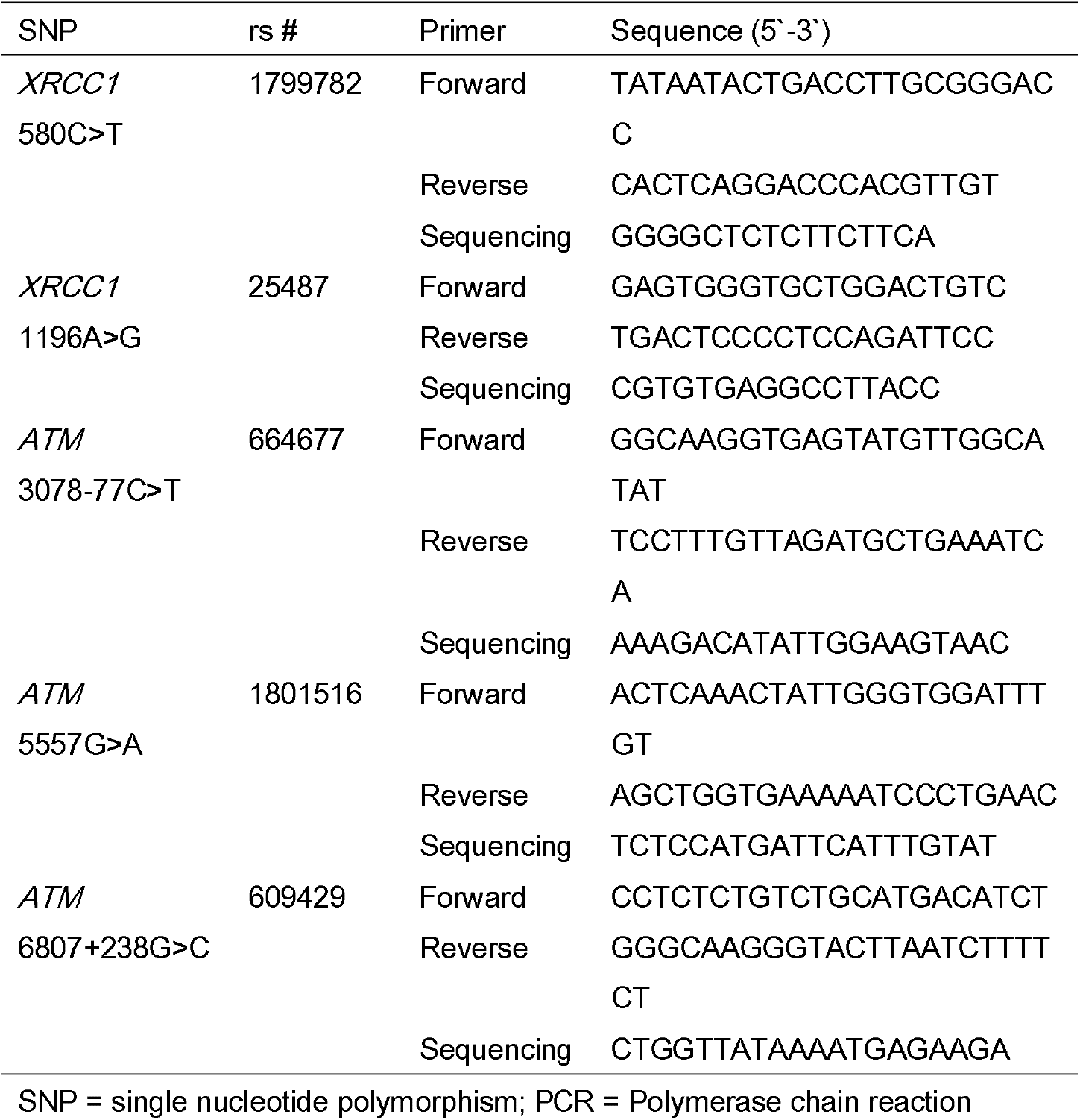
Characteristics of SNP and PCR primers

Each biotinylated PCR product (20μl) was denatured and immobilized on Streptavidin Sepharose High Performance beads (Amersham Bioscience, Piscataway, NJ, USA) using the PSQ Vacuum Prep Tool (Qiagen, Hilden, Germany) and Pyromark Q96 Vacuum Prep Worktable (Qiagen, Hilden, Germany). These were then incubated at 72-80°C for 3 minutes with 10pM of sequencing primer and placed in the room temperature. Pyrosequencing was performed using PyroGold Q96 Reagents (Qiagen, Hilden, Germany) on the Pyromark Q96 ID instrument (Qiagen, Hilden, Germany) according to the manufacturer’s instructions. Pyrogram was analyzed by the Pyromark software version 1.0 (Qiagen, Hilden, Germany)(Figure 1). When the result of pyrogram analysis was indeterminate, pyrosequencing and subsequent analyses were repeated two times. If the result of analysis was still indeterminate, the genotype was determined as genotyping failure.

**Figure 1.**
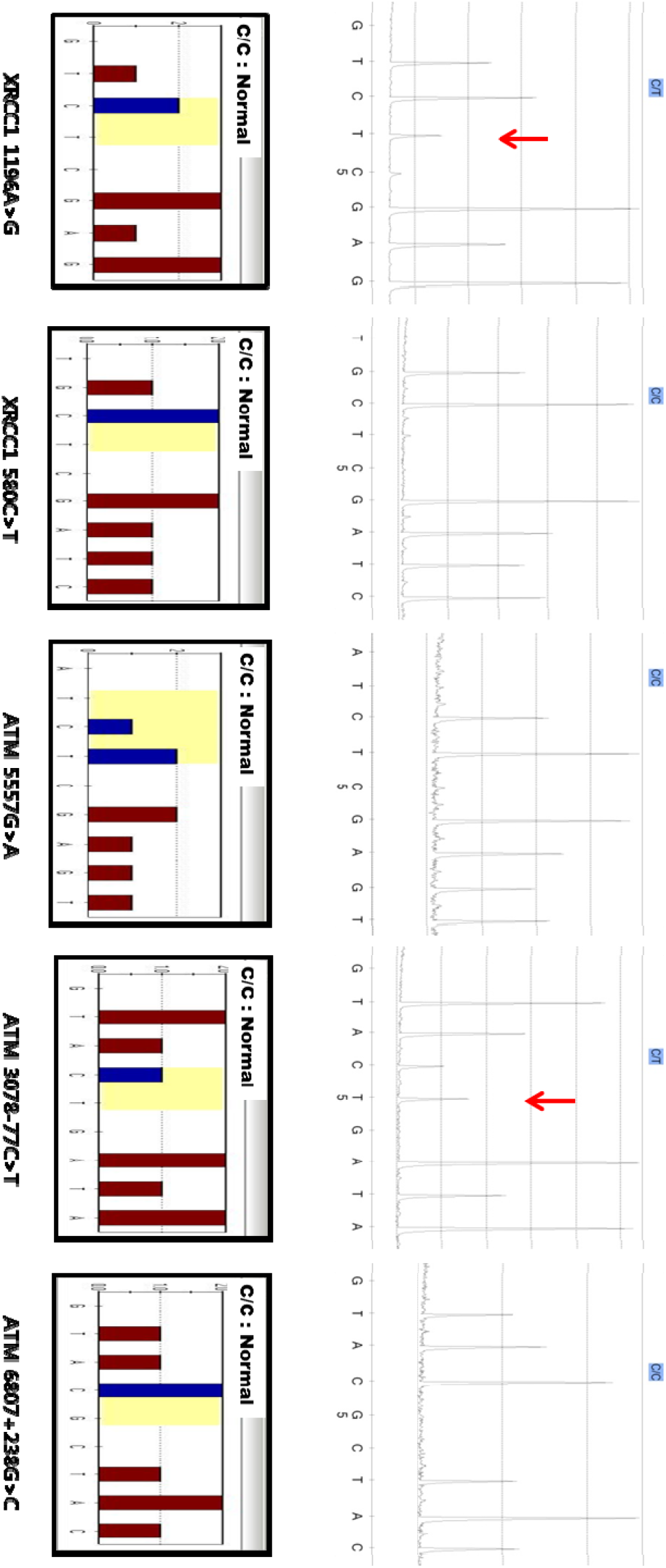
Example of the pyrogram analysis. Arrows indicate the existence of minor alleles.

### Statistical analysis

Clinicopathologic variables and genotype of SNPs were summarized. Hardy-Weinberg equilibrium was tested using chi-square test. The concordance rate of genotype from cancer tissue and that from normal tissue was calculated according to SNP. To evaluate the association of clinicopathologic variables with RFS and OS, all clinicopathologic variables except risk group were dichotomized using the median or appropriate value. The association of clinicopathologic variables with RFS and OS was evaluated via Cox regression analysis. Forward stepwise selection method was used to select clinicopathologic variables independently associated with RFS or OS. The association of genotype of SNPs with RFS and OS was evaluated via Cox regression analysis under the adjustment for clinicopathologic variables independently associated with RFS or OS. The association of genotype of SNPs with completion of CRT was evaluated via chi-square or Fisher’s exact test. Haplotype estimation was performed using haplo.stats in R version 2.15.0 (R Development Core Team (2012). R: A language and environment for statistical computing, reference index version 2.15.0. R Foundation for Statistical Computing, Vienna, Austria. ISBN 3-900051-07-0, URL http://www.R-project.org.). A P-value<0.05 was regarded as statistically significant and all statistical analyses except haplotype estimation were performed using SPSS (version 18.0) (SPSS Inc., Chicago, IL, USA).

## Results

### Baseline characteristics

The baseline characteristics of 153 patients are summarized in Table 2. The median age was 47 years and median clinical tumor size was 3.0 cm. Stage 1B1 accounted for half of population and squamous cell carcinoma is the most common histologic type. The median pathologic tumor size was 3.3 cm and invasion to outer 1/3 of cervical wall was present in 69% of patients. Lymphovascular space invasion (LVSI), parametrial invasion, positive resection margin and lymph node involvement were present in 56%, 25%, 7% and 44% of patients, respectively. Over half of patients were classified as high-risk and 16 of 153 patients could not complete the adjuvant CRT. During follow-up, 29 recurrences and deaths were observed (Table 2). Specifically, one vaginal, three pelvic and 25 extra-pelvic recurrences have occurred.

**Table 2.** Baseline characteristics

| Characteristics | n | (%) |
| --- | --- | --- |
| Age, year, median (range) | 47 | (25-69) |
| Clinical tumor size, cm, median (range) | 3.0 | (0.5-6.0) |
| Stage |  |  |
| 1B1 | 81 | (53) |
| 1B2 | 21 | (14) |
| 2A1 | 31 | (20) |
| 2A2 | 20 | (13) |
| Histologic type |  |  |
| Squamous | 124 | (81) |
| Non-squamous | 29 | (19) |
| Pathologic tumor size, cm, median (range) | 3.3 | (0.7-7.5) |
| Invasion depth |  |  |
| Inner 1/3 | 6 | (4) |
| Middle 1/3 | 41 | (27) |
| Outer 1/3 | 106 | (69) |
| LVSI |  |  |
| Yes | 85 | (56) |
| No | 68 | (44) |
| Parametrial invasion |  |  |
| Yes | 38 | (25) |
| No | 115 | (75) |
| Resection margin |  |  |
| Positive | 11 | (7) |
| Negative | 142 | (93) |
| Lymph node involvement |  |  |
| Yes | 68 | (44) |
| No | 85 | (56) |
| Risk group |  |  |
| Low | 29 | (19) |
| Intermediate | 38 | (25) |
| High | 86 | (56) |
| Completion of CRT |  |  |
| Yes | 137 | (90) |
| No | 16 | (10) |
| Recurrence | 29 | (19) |
| Death | 29 | (19) |

LVSI = lymphovascular space invasion; CRT = concurrent chemoradiation

### Genotype of SNP

All SNPs except *ATM* 6807+23G>C from cancer tissue satisfied the Hardy-Weinberg equilibrium. The frequency of minor genotypes was considerable except *ATM* 5557G>A. Genotyping failure occurred more frequently in normal tissue (7%) than cancer tissue (1%). In addition, genotyping failure occurred more frequently in *ATM* 6807+238G>C (14%) than other SNPs (1%)(Table 3).

**Table 3.**
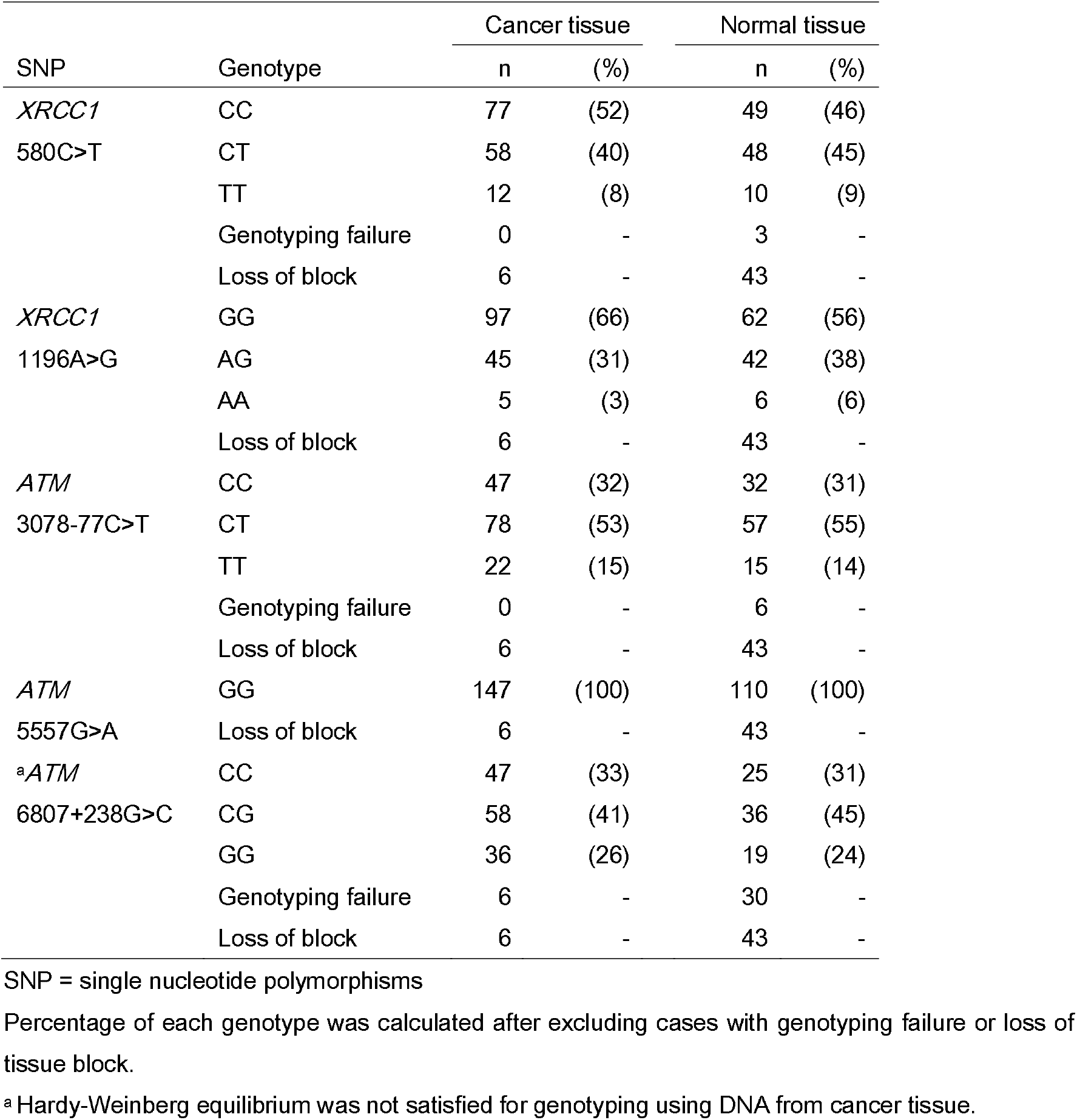
Genotype of single nucleotide polymorphism

| SNP | Genotype | Cancer tissue |  | Normal tissue |  |
| --- | --- | --- | --- | --- | --- |
|  |  | n | (%) | n | (%) |
| *XRCC1* | CC | 77 | (52) | 49 | (46) |
| 580C>T | CT | 58 | (40) | 48 | (45) |
|  | TT | 12 | (8) | 10 | (9) |
|  | Genotyping failure | 0 | - | 3 | - |
|  | Loss of block | 6 | - | 43 | - |
| *XRCC1* | GG | 97 | (66) | 62 | (56) |
| 1196A>G | AG | 45 | (31) | 42 | (38) |
|  | AA | 5 | (3) | 6 | (6) |
|  | Loss of block | 6 | - | 43 | - |
| *ATM* | CC | 47 | (32) | 32 | (31) |
| 3078-77C>T | CT | 78 | (53) | 57 | (55) |
|  | TT | 22 | (15) | 15 | (14) |
|  | Genotyping failure | 0 | - | 6 | - |
|  | Loss of block | 6 | - | 43 | - |
| *ATM* | GG | 147 | (100) | 110 | (100) |
| 5557G>A | Loss of block | 6 | - | 43 | - |
| <sup>a</sup>*ATM* | CC | 47 | (33) | 25 | (31) |
| 6807+238G>C | CG | 58 | (41) | 36 | (45) |
|  | GG | 36 | (26) | 19 | (24) |
|  | Genotyping failure | 6 | - | 30 | - |
|  | Loss of block | 6 | - | 43 | - |

SNP = single nucleotide polymorphisms
Percentage of each genotype was calculated after excluding cases with genotyping failure or loss of tissue block.
<sup>a</sup> Hardy-Weinberg equilibrium was not satisfied for genotyping using DNA from cancer tissue.

### Concordance between genotype from cancer tissue and that from normal tissue

Concordance rate varied according to SNP. Specifically, concordance rates of *XRCC1* 580C>T, *XRCC1* 1196A>G, *ATM* 3078-77C>T, *ATM* 5557G>A and *ATM* 6807+238G>C were 72% (Table 4), 88% (Table 5), 73% (Table 6), 100% and 54% (Table 7), respectively.

**Table 4.**
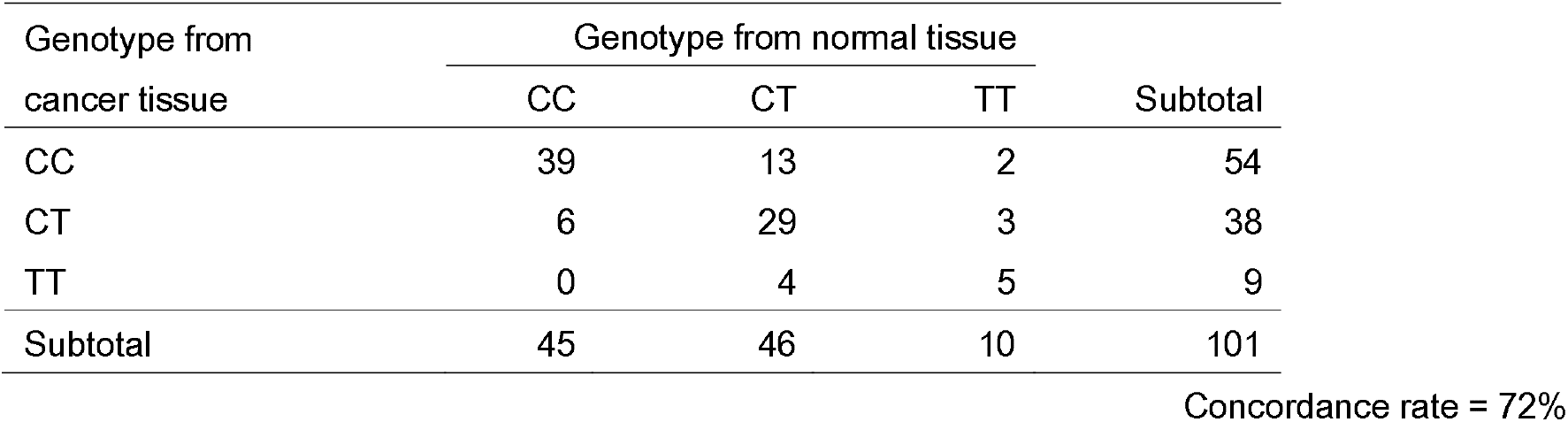
Concordance between genotype from cancer tissue and that from normal tissue for *XRCC1* 580C>T

**Table 5.** Concordance between genotype from cancer tissue and that from normal tissue for *XRCC1* 1196A>G

| Genotype from<br>cancer tissue | Genotype from normal tissue |  |  | Subtotal |
| --- | --- | --- | --- | --- |
|  | GG | AG | AA |  |
| GG | 58 | 9 | 1 | 68 |
| AG | 1 | 30 | 2 | 33 |
| AA | 0 | 0 | 3 | 3 |
| Subtotal | 59 | 39 | 6 | 104 |

Concordance rate = 88%

**Table 6.**
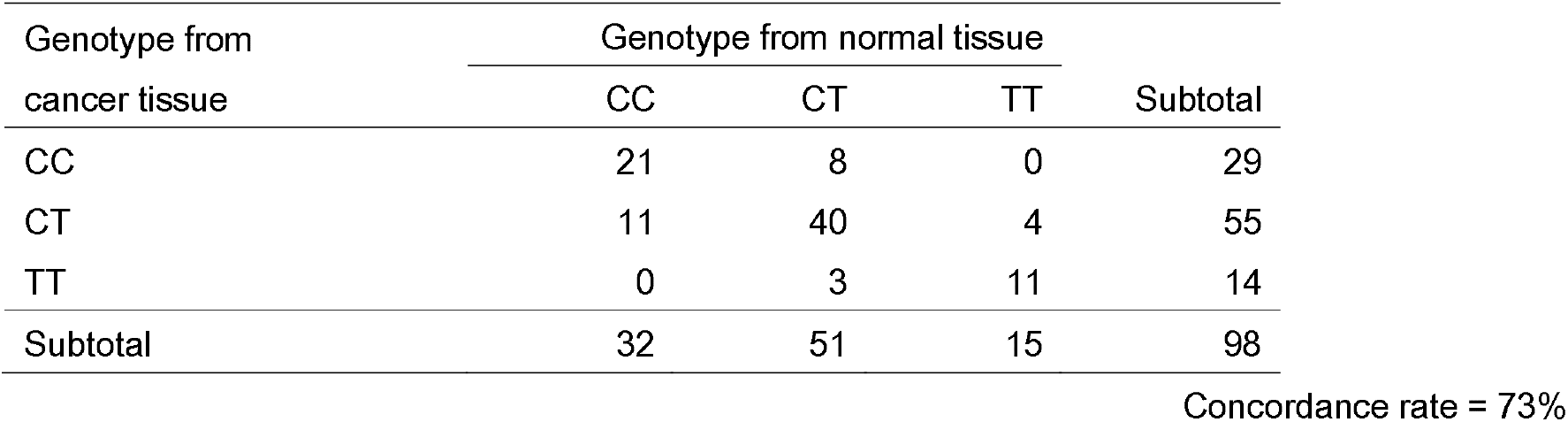
Concordance between genotype from cancer tissue and that from normal tissue for *ATM* 3078-77C>T

| Genotype from<br>cancer tissue | Genotype from normal tissue |  |  | Subtotal |
| --- | --- | --- | --- | --- |
|  | CC | CT | TT |  |
| CC | 21 | 8 | 0 | 29 |
| CT | 11 | 40 | 4 | 55 |
| TT | 0 | 3 | 11 | 14 |
| Subtotal | 32 | 51 | 15 | 98 |

Concordance rate = 73%

**Table 7.** Concordance between genotype from cancer tissue and that from normal tissue for *ATM* 6807+238G>C

| Genotype from<br>cancer tissue | Genotype from normal tissue |  |  | Subtotal |
| --- | --- | --- | --- | --- |
|  | CC | CT | TT |  |
| CC | 14 | 8 | 2 | 24 |
| CT | 8 | 16 | 7 | 31 |
| TT | 2 | 7 | 10 | 19 |
| Subtotal | 24 | 31 | 19 | 74 |

Concordance rate = 54%

### Association of clinicopathologic variables with RFS and OS

In univariate analysis, age, clinical tumor size, stage, histologic type, pathologic tumor size, LVSI, parametrial invasion, resection margin, lymph node involvement and risk group were associated with RFS. In multivariate analysis, age, stage, histologic type, LVSI and parametrial invasion were associated with RFS (Table 8).

**Table 8.**
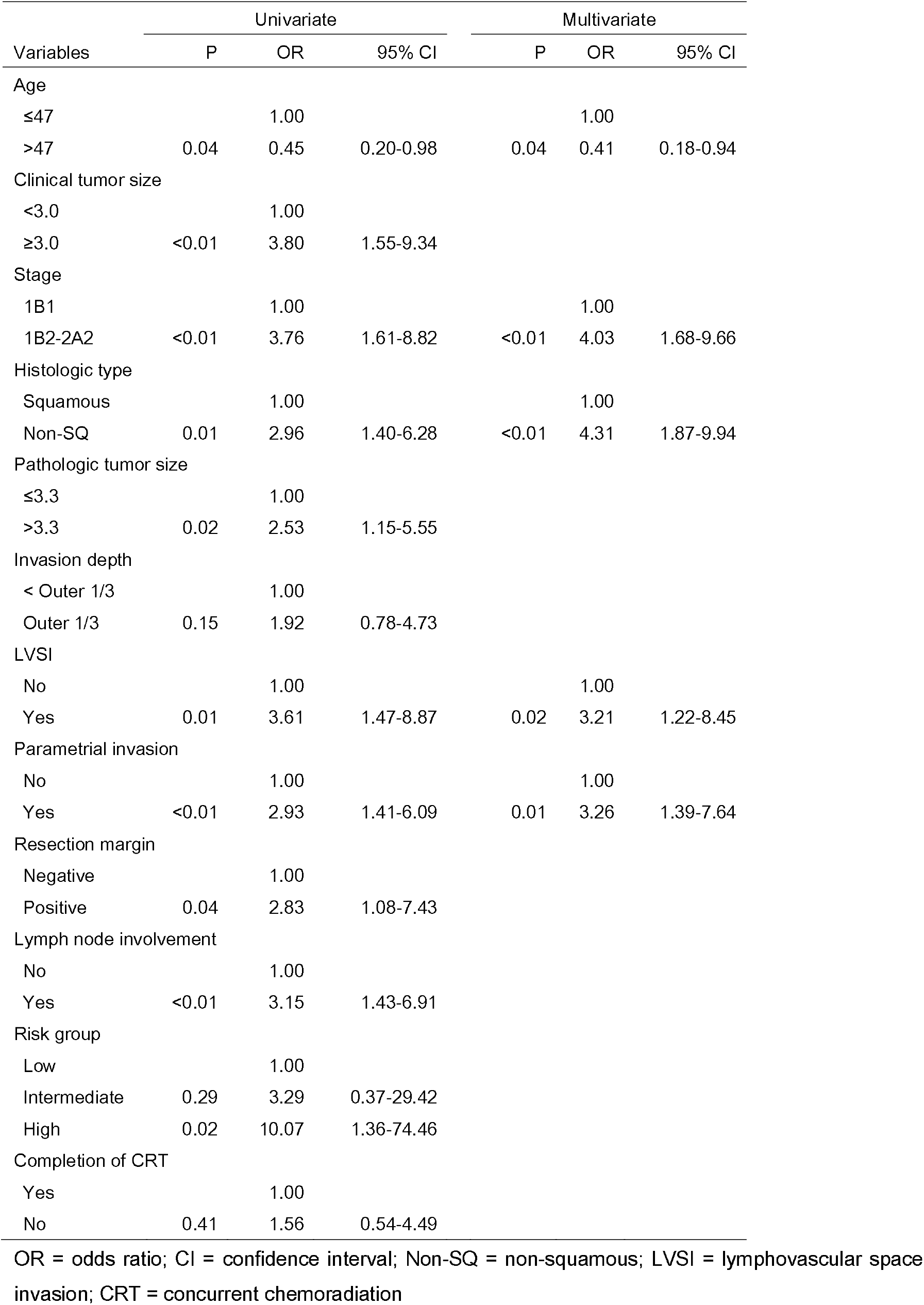
Association of clinicopathologic variables with recurrence-free survival

| Variables | Univariate |  |  | Multivariate |  |  |
| --- | --- | --- | --- | --- | --- | --- |
|  | P | OR | 95% CI | P | OR | 95% CI |
| Age |  |  |  |  |  |  |
| ≤47 |  | 1.00 |  |  | 1.00 |  |
| >47 | 0.04 | 0.45 | 0.20-0.98 | 0.04 | 0.41 | 0.18-0.94 |
| Clinical tumor size |  |  |  |  |  |  |
| <3.0 |  | 1.00 |  |  |  |  |
| ≥3.0 | <0.01 | 3.80 | 1.55-9.34 |  |  |  |
| Stage |  |  |  |  |  |  |
| 1B1 |  | 1.00 |  |  | 1.00 |  |
| 1B2-2A2 | <0.01 | 3.76 | 1.61-8.82 | <0.01 | 4.03 | 1.68-9.66 |
| Histologic type |  |  |  |  |  |  |
| Squamous |  | 1.00 |  |  | 1.00 |  |
| Non-SQ | 0.01 | 2.96 | 1.40-6.28 | <0.01 | 4.31 | 1.87-9.94 |
| Pathologic tumor size |  |  |  |  |  |  |
| ≤3.3 |  | 1.00 |  |  |  |  |
| >3.3 | 0.02 | 2.53 | 1.15-5.55 |  |  |  |
| Invasion depth |  |  |  |  |  |  |
| < Outer 1/3 |  | 1.00 |  |  |  |  |
| Outer 1/3 | 0.15 | 1.92 | 0.78-4.73 |  |  |  |
| LVSI |  |  |  |  |  |  |
| No |  | 1.00 |  |  | 1.00 |  |
| Yes | 0.01 | 3.61 | 1.47-8.87 | 0.02 | 3.21 | 1.22-8.45 |
| Parametrial invasion |  |  |  |  |  |  |
| No |  | 1.00 |  |  | 1.00 |  |
| Yes | <0.01 | 2.93 | 1.41-6.09 | 0.01 | 3.26 | 1.39-7.64 |
| Resection margin |  |  |  |  |  |  |
| Negative |  | 1.00 |  |  |  |  |
| Positive | 0.04 | 2.83 | 1.08-7.43 |  |  |  |
| Lymph node involvement |  |  |  |  |  |  |
| No |  | 1.00 |  |  |  |  |
| Yes | <0.01 | 3.15 | 1.43-6.91 |  |  |  |
| Risk group |  |  |  |  |  |  |
| Low |  | 1.00 |  |  |  |  |
| Intermediate | 0.29 | 3.29 | 0.37-29.42 |  |  |  |
| High | 0.02 | 10.07 | 1.36-74.46 |  |  |  |
| Completion of CRT |  |  |  |  |  |  |
| Yes |  | 1.00 |  |  |  |  |
| No | 0.41 | 1.56 | 0.54-4.49 |  |  |  |

OR = odds ratio; CI = confidence interval; Non-SQ = non-squamous; LVSI = lymphovascular space invasion; CRT = concurrent chemoradiation

In univariate analysis, clinical tumor size, histologic type, LVSI, parametrial invasion, resection margin, lymph node involvement, risk group and completion of CRT were associated with OS. In multivariate analysis, clinical tumor size, resection margin, lymph node involvement and completion of CRT were associated with OS (Table 9).

**Table 9.** Association of clinicopathologic variables with overall survival

| Variables | Univariate |  |  | Multivariate |  |  |
| --- | --- | --- | --- | --- | --- | --- |
|  | P | OR | 95% CI | P | OR | 95% CI |
| Age |  |  |  |  |  |  |
| ≤47 |  | 1.00 |  |  |  |  |
| >47 | 0.20 | 0.61 | 0.29-1.29 |  |  |  |
| Clinical tumor size |  |  |  |  |  |  |
| <3.0 |  | 1.00 |  |  | 1.00 |  |
| ≥3.0 | 0.04 | 2.38 | 1.05-5.37 | 0.02 | 2.58 | 1.14-5.88 |
| Stage |  |  |  |  |  |  |
| 1B1 |  | 1.00 |  |  |  |  |
| 1B2-2A2 | 0.08 | 1.97 | 0.93-4.17 |  |  |  |
| Histologic type |  |  |  |  |  |  |
| Squamous |  | 1.00 |  |  |  |  |
| Non-SQ | 0.01 | 2.72 | 1.26-5.85 |  |  |  |
| Pathologic tumor size |  |  |  |  |  |  |
| ≤3.3 |  | 1.00 |  |  |  |  |
| >3.3 | 0.26 | 1.53 | 0.73-3.20 |  |  |  |
| Invasion depth |  |  |  |  |  |  |
| < Outer 1/3 |  | 1.00 |  |  |  |  |
| Outer 1/3 | 0.24 | 1.72 | 0.70-4.22 |  |  |  |
| LVSI |  |  |  |  |  |  |
| No |  | 1.00 |  |  |  |  |
| Yes | 0.02 | 2.83 | 1.21-6.64 |  |  |  |
| Parametrial invasion |  |  |  |  |  |  |
| No |  | 1.00 |  |  |  |  |
| Yes | 0.02 | 2.34 | 1.12-4.90 |  |  |  |
| Resection margin |  |  |  |  |  |  |
| Negative |  | 1.00 |  |  | 1.00 |  |
| Positive | <0.01 | 3.96 | 1.61-9.75 | 0.01 | 3.70 | 1.49-9.19 |
| Lymph node involvement |  |  |  |  |  |  |
| No |  | 1.00 |  |  | 1.00 |  |
| Yes | <0.01 | 3.27 | 1.49-7.18 | <0.01 | 3.18 | 1.44-7.04 |
| Risk group |  |  |  |  |  |  |
| Low |  | 1.00 |  |  |  |  |
| Intermediate | 0.15 | 4.66 | 0.56-38.72 |  |  |  |
| High | 0.04 | 8.53 | 1.15-63.33 |  |  |  |
| Completion of CRT |  |  |  |  |  |  |
| Yes |  | 1.00 |  |  | 1.00 |  |
| No | 0.02 | 2.88 | 1.17-7.10 | 0.02 | 3.10 | 1.24-7.75 |

OR = odds ratio; CI = confidence interval; Non-SQ = non-squamous; LVSI = lymphovascular space invasion; CRT = concurrent chemoradiation

### Adjusted RFS and OS according to genotype

Genotype of *XRCC1* 580C>T was not associated with RFS in patients with cervical cancer who underwent radical surgery followed by adjuvant CRT (Figure 2). Of all SNPs, genotype from cancer tissue was associated with neither RFS nor OS adjusted for clinicopathologic variables (Table 10). Similarly, genotype from normal tissue was associated with neither RFS nor OS adjusted for clinicopathologic variables (Table 11).

**Table 10.**
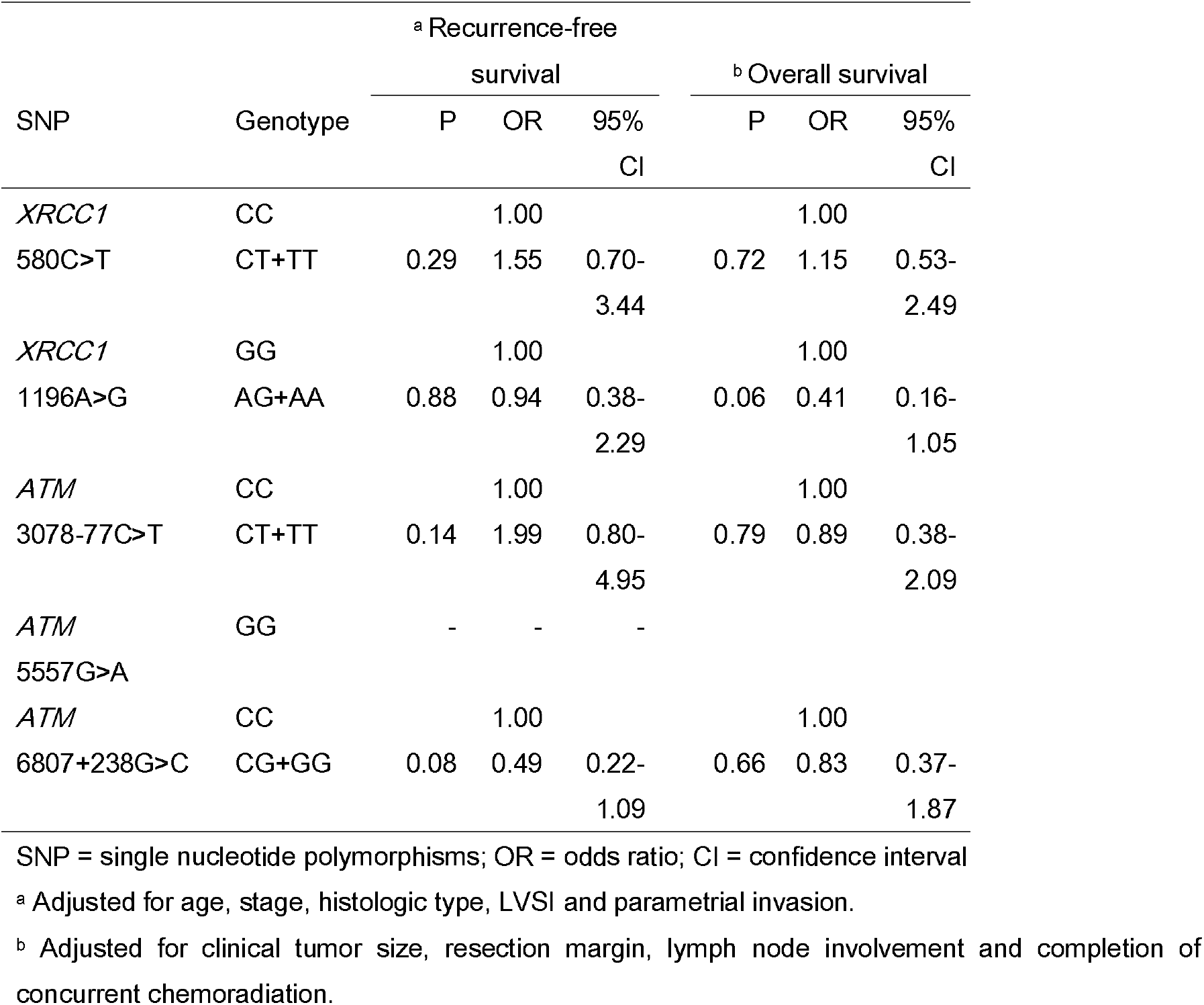
Association of genotype from cancer tissue with recurrence-free and overall survival

| SNP | Genotype | <sup>a</sup> Recurrence-free survival |  |  | <sup>b</sup> Overall survival |  |  |
| --- | --- | --- | --- | --- | --- | --- | --- |
|  |  | P | OR | 95% CI | P | OR | 95% CI |
| *XRCC1* 580C>T | CC |  | 1.00 |  |  | 1.00 |  |
|  | CT+TT | 0.29 | 1.55 | 0.70-3.44 | 0.72 | 1.15 | 0.53-2.49 |
| *XRCC1* 1196A>G | GG |  | 1.00 |  |  | 1.00 |  |
|  | AG+AA | 0.88 | 0.94 | 0.38-2.29 | 0.06 | 0.41 | 0.16-1.05 |
| *ATM* 3078-77C>T | CC |  | 1.00 |  |  | 1.00 |  |
|  | CT+TT | 0.14 | 1.99 | 0.80-4.95 | 0.79 | 0.89 | 0.38-2.09 |
| *ATM* 5557G>A | GG | - | - | - |  |  |  |
| *ATM* 6807+238G>C | CC |  | 1.00 |  |  | 1.00 |  |
|  | CG+GG | 0.08 | 0.49 | 0.22-1.09 | 0.66 | 0.83 | 0.37-1.87 |

SNP = single nucleotide polymorphisms; OR = odds ratio; CI = confidence interval
<sup>a</sup> Adjusted for age, stage, histologic type, LVSI and parametrial invasion.
<sup>b</sup> Adjusted for clinical tumor size, resection margin, lymph node involvement and completion of concurrent chemoradiation.

**Table 11.** Association of genotype from normal tissue with recurrence-free and overall survival

| SNP | Genotype | <sup>a</sup> Recurrence-free survival |  |  | <sup>b</sup> Overall survival |  |  |
| --- | --- | --- | --- | --- | --- | --- | --- |
|  |  | P | OR | 95% CI | P | OR | 95% CI |
| *XRCC1* 580C>T | CC |  | 1.00 |  |  | 1.00 |  |
|  | CT+TT | 0.26 | 1.68 | 0.68-4.18 | 0.29 | 1.64 | 0.65-4.15 |
| *XRCC1* 1196A>G | GG |  | 1.00 |  |  | 1.00 |  |
|  | AG+AA | 0.78 | 0.89 | 0.37-2.10 | 0.27 | 0.62 | 0.26-1.46 |
| *ATM* 3078-77C>T | CC |  | 1.00 |  |  | 1.00 |  |
|  | CT+TT | 0.38 | 1.64 | 0.54-4.98 | 0.15 | 2.32 | 0.75-7.16 |
| *ATM* 5557G>A | GG | - | - | - |  |  |  |
| *ATM* 6807+238G>C | CC |  | 1.00 |  |  | 1.00 |  |
|  | CG+GG | 0.16 | 2.50 | 0.69-9.03 | 0.25 | 2.52 | 0.53-12.02 |

SNP = single nucleotide polymorphisms; OR = odds ratio; CI = confidence interval
<sup>a</sup> Adjusted for age, stage, histologic type, LVSI and parametrial invasion.
<sup>b</sup> Adjusted for clinical tumor size, resection margin, lymph node involvement and completion of concurrent chemoradiation.

**Figure 2.**
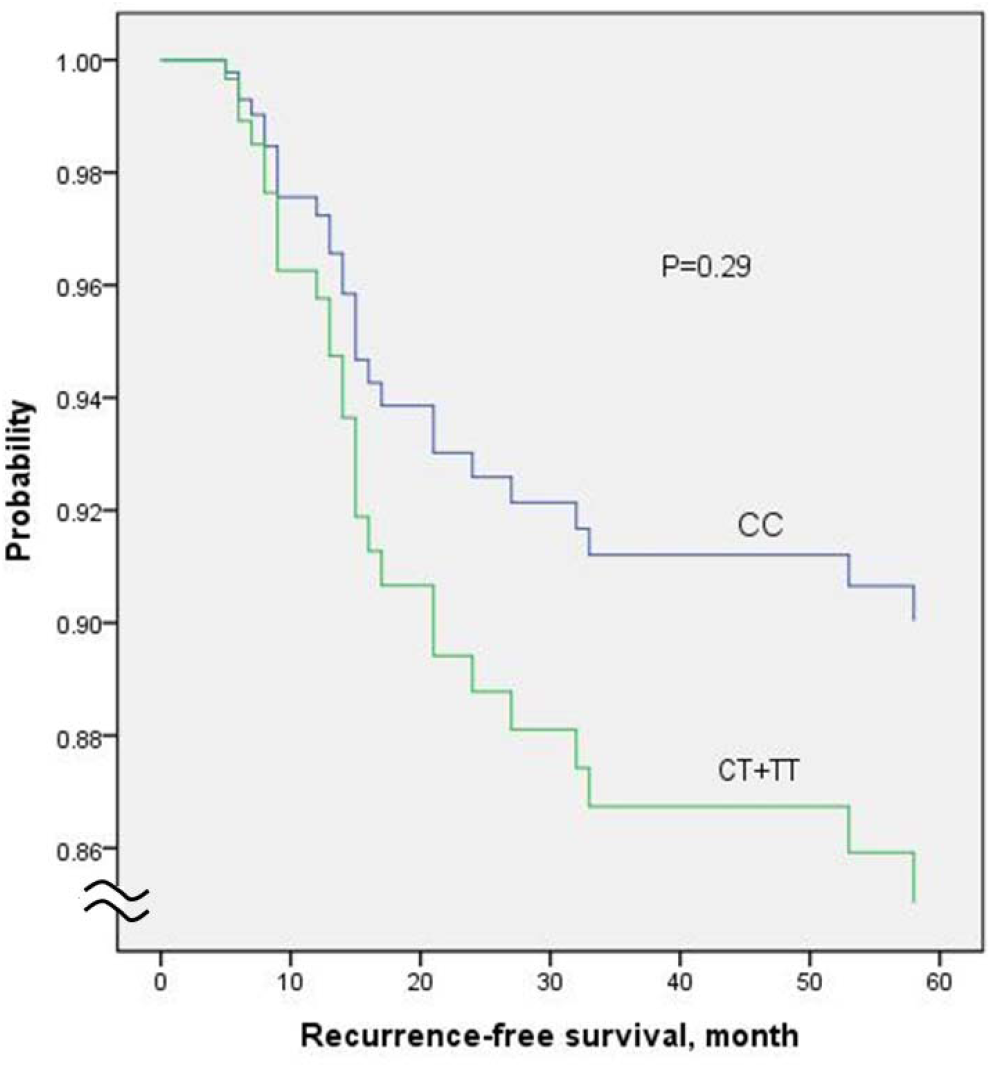
Recurrence-free survival according to genotype of *XRCC1* 580C>T

### Association of haplotype with RFS and OS

The distribution of haplotypes among *XRCC1* and *ATM* SNPs was summarized in Table 12 and Table 13, respectively. Haplotype pairs in *XRCC1* were associated with neither RFS nor OS (Table 14). Similarly, haplotype pairs in *ATM* were associated with neither RFS nor OS (Table 15).

**Table 12.** Distribution of haplotypes among *XRCC1* polymorphisms

| <sup>a</sup> Haplotype | Portion of haplotype in<br>cancer tissue | Portion of haplotype<br>in normal tissue |
| --- | --- | --- |
| CG | 0.54 | 0.46 |
| TG | 0.27 | 0.29 |
| CA | 0.18 | 0.22 |
| TA | 0.01 | 0.03 |

<sup>a</sup> Composed of two polymorphic sites: *XRCC1* 580C>T, *XRCC1* 1196A>G.

**Table 13.**
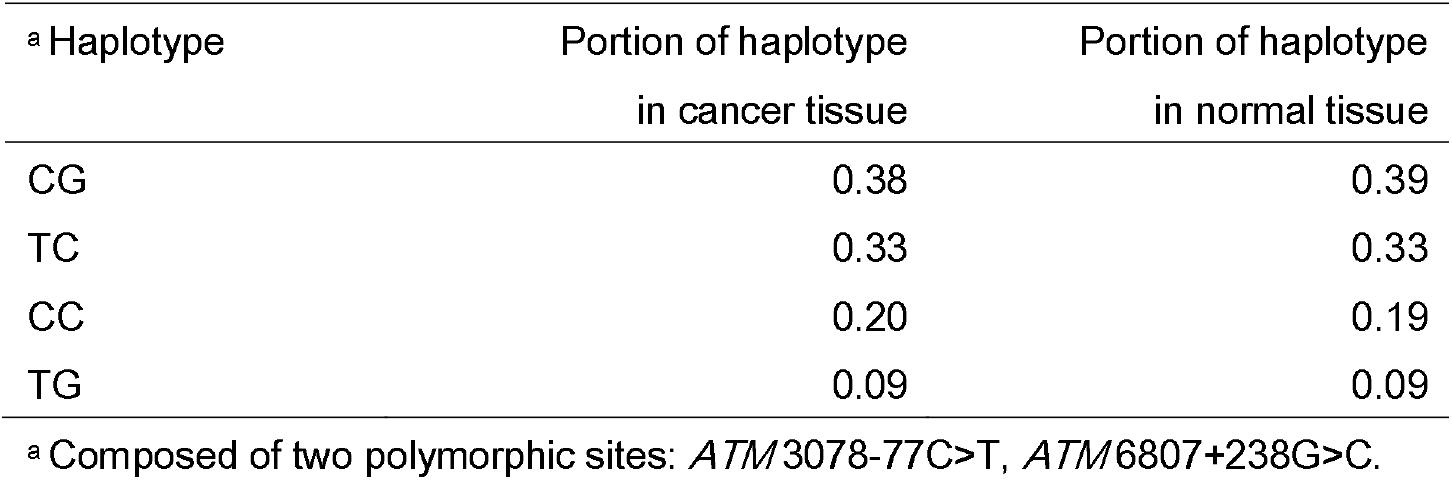
Distribution of haplotypes among *ATM* polymorphisms

**Table 14.** Association of haplotype pairs of *XRCC1* with RFS and OS

| Haplotype pairs | <sup>a</sup> Recurrence-free survival |  |  | <sup>b</sup> Overall survival |  |  |
| --- | --- | --- | --- | --- | --- | --- |
|  | P | OR | 95% CI | P | OR | 95% CI |
| Cancer tissue |  |  |  |  |  |  |
| CG/CG |  | 1.00 |  |  | 1.00 |  |
| CG/Others | 0.99 | 0.99 | 0.42-2.38 | 0.97 | 0.98 | 0.42-2.29 |
| Others/Others | 0.60 | 1.29 | 0.50-3.36 | 0.85 | 1.10 | 0.42-2.89 |
| Normal tissue |  |  |  |  |  |  |
| CG/CG |  | 1.00 |  |  | 1.00 |  |
| CG/Others | 0.92 | 1.04 | 0.49-2.22 | 0.42 | 1.39 | 0.63-3.09 |
| Others/Others | 0.85 | 1.09 | 0.48-2.47 | 0.73 | 0.85 | 0.34-2.12 |

OR = odds ratio; CI = confidence interval
<sup>a</sup> Adjusted for age, stage, histologic type, LVSI and parametrial invasion.
<sup>b</sup> Adjusted for clinical tumor size, resection margin, lymph node involvement and completion of concurrent chemoradiation.

**Table 15.** Association of haplotype pairs of *ATM* with RFS and OS

| Haplotype pairs | <sup>a</sup> Recurrence-free survival |  |  | <sup>b</sup> Overall survival |  |  |
| --- | --- | --- | --- | --- | --- | --- |
|  | P | OR | 95% CI | P | OR | 95% CI |
| Cancer tissue |  |  |  |  |  |  |
| CG/CG |  | 1.00 |  |  | 1.00 |  |
| CG/Others | 0.27 | 1.81 | 0.64-5.16 | 0.37 | 1.57 | 0.59-4.18 |
| Others/Others | 0.45 | 1.51 | 0.52-4.42 | 0.98 | 0.99 | 0.35-2.78 |
| Normal tissue |  |  |  |  |  |  |
| CG/CG |  | 1.00 |  |  | 1.00 |  |
| CG/Others | 0.18 | 1.85 | 0.75-4.60 | 0.20 | 1.82 | 0.73-4.53 |
| Others/Others | 0.19 | 1.88 | 0.73-4.79 | 0.38 | 1.53 | 0.59-3.96 |

OR = odds ratio; CI = confidence interval
<sup>a</sup> Adjusted for age, stage, histologic type, LVSI and parametrial invasion.
<sup>b</sup> Adjusted for clinical tumor size, resection margin, lymph node involvement and completion of concurrent chemoradiation.

### Association of SNPs with completion of CRT

In the SNPs, genotype from cancer tissue was not associated with completion of CRT (Table 16). Similarly, genotype from normal tissue was not associated with completion of CRT (Table 17).

**Table 16.** Association of genotype from cancer tissue with completion of CRT

| SNP | Genotype | Completion of CRT |  | P value |
| --- | --- | --- | --- | --- |
|  |  | No | Yes |  |
| *XRCC1* 580C>T | CC | 9 | 68 | 0.74 |
|  | CT+TT | 7 | 63 |  |
| *XRCC1* 1196A>G | GG | 8 | 89 | 0.15 |
|  | AG+AA | 8 | 42 |  |
| *ATM* 3078-77C>T | CC | 3 | 44 | 0.23 |
|  | CT+TT | 13 | 87 |  |
| *ATM* 5557G>A | GG | 16 | 131 | - |
| *ATM* 6807+238G>C | CC | 5 | 42 | 0.85 |
|  | CG+GG | 11 | 83 |  |

SNP = single nucleotide polymorphisms; CRT = concurrent chemoradiation

**Table 17.**
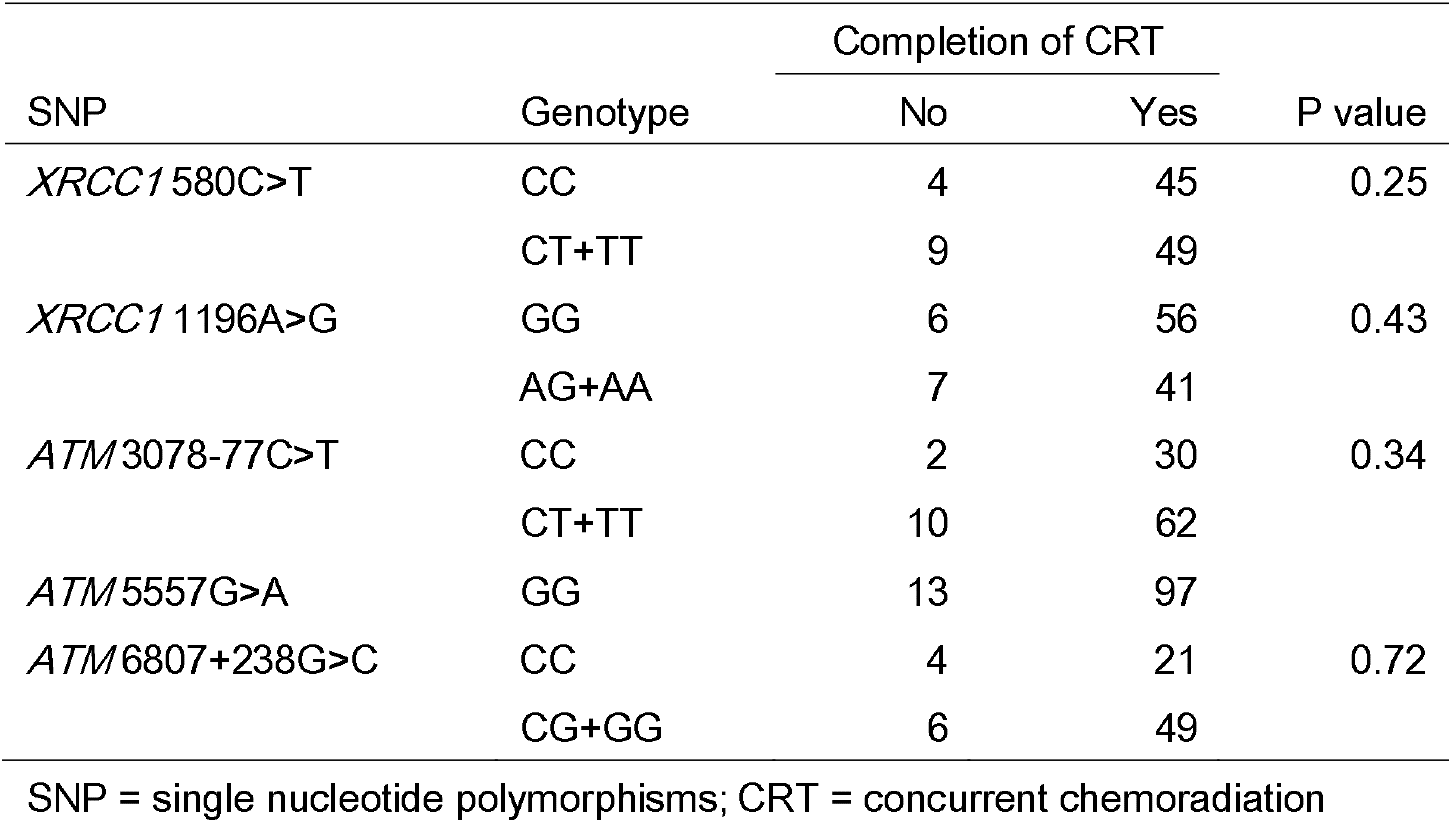
Association of genotype from normal tissue with completion of CRT

## Discussion

### RFS according to *XRCC1* 580C>T

This study failed to show different RFS according to genotype of *XRCC1* 580C>T in patients with cervical cancer who underwent radical surgery followed by adjuvant CRT. There was a trend that patients with CT or TT genotype had a worse RFS than those with CC genotype but the difference was not significant.

I believe that inadequate sample size as a result of lower than expected effect is the main reason for the negative result. Specifically, I calculated the sample size using estimated 2-year RFS of 27 patients included in my previous study.^8^ The estimated 2-year RFS of patients with CC genotype was 85.9% and that of patients with CT or TT genotype was 63.6%. However, in this study, the estimated 2-year RFS of patients with CC genotype was 87.4% and that of patients with CT or TT genotype was 80.9%. The incorrect estimation of 2-year RFS in patients with CT or TT genotype resulted in small sample size and lack of power to detect the difference in RFS. I recalculated the sample size using the observed 2-year RFS in the current study. To achieve statistical power of at least 0.80 when keeping the probability of type 1 error at 0.05, 550 patients per arm is necessary considering the 10% genotyping failure rate, which is about eight-fold larger than initially calculated sample size (n=70 per arm).

### Concordance between genotype from cancer tissue and that from normal tissue

Various small studies of colon and breast cancer reported high concordance rate between genotype from cancer tissue and that from normal tissue.^15-19^ For example, in a study including 44 colorectal cancer samples and adjacent normal tissue identified strong concordance between paired samples for 28 SNPs in 13 genes and the concordance rate ranged between 92% and 100%.^16^ However, the current study showed quite a low concordance rate ranging between 54% and 100%.

The current study suggested that the variability between cancer and germline genotype at the SNP level could be significant in cervical cancer and the germline genome could not be used as a surrogate to assess SNP status in the cancer.

### Association of SNPs in *XRCC1* and *ATM* with RFS and OS

Whether DNA is extracted from cancer tissue or normal tissue, none of SNPs were associated with RFS or OS adjusted for clinicopathologic variables. In addition, none of haplotypes of SNPs was associated with RFS or OS. I believe that lack of power due to small sample size is one of the reasons for negative results.

### Association of clinicopathologic variables with RFS and OS

Patients with high-risk factors such as parametrial invasion, positive resection margin and lymph node involvement were known to be at high-risk for recurrence and adjuvant treatment was indicated.2 Similarly, patients with intermediate-risk factors were also known to have modestly increased risk for recurrence and usually received adjuvant treatment.^3,5^ However, there is scant literature regarding prognostic factors in patients who already received postoperative adjuvant treatment. Specifically, it is unclear whether the increased risk of patients with high- or intermediate-risk factors is negated by adjuvant treatment.

The current study showed that the recurrence risk of patients with age≤47, stage>1B1, non-squamous histologic type, LVSI or parametrial invasion is still elevated despite of adjuvant treatment and demonstrated that death risk of patients with large tumor, positive resection margin, lymph node involvement or failure to complete CRT is incompletely negated by adjuvant CRT. Patients with such risk factors could be the target population for additional treatment after radical surgery followed by adjuvant CRT.

### Genotyping failure

If high quality DNA from lymphocyte or fresh frozen tissues is used, the genotype of SNP can be accurately determined in most cases. However, the quality of DNA extracted from paraffin blocks is known to be inferior to DNA from lymphocyte or fresh frozen tissues.^20^ Therefore, SNP genotyping using DNA from paraffin blocks has lower call rates and accuracy compared to genotyping using fresh frozen tissues.^20^ Reduced DNA extraction efficiency, reduced quality due to cross-linking and degradation, fragmentation of DNA and selective non-amplification of large fragments, inaccurate SNP calls in GC rich regions were suggested as the mechanisms of low performance of genotyping using DNA from paraffin blocks.^20^

In the current study, genotyping failure occurred more frequently in normal tissue (7%) than cancer tissue (1%). The mechanism for more genotyping failure in normal tissue than in cancer tissue is unclear. In addition, genotyping failure occurred more frequently in *ATM* 6807+238G>C (14%) than other SNPs (1%). The large size of the amplicon of *ATM* 6807+238G>C could be the reason for frequent genotyping failure. The size and GC contents (%) of amplicon of each SNP were summarized in Table 18.

**Table 18.** Size and GC contents (%) of amplicon

|  | Size (bp) | GC contents (%) |
| --- | --- | --- |
| *XRCC1* 580C>T | 168 | 56 |
| *XRCC1* 1196A>G | 120 | 63 |
| *ATM* 3078-77C>T | 169 | 33 |
| *ATM* 5557G>A | 158 | 34 |
| *ATM* 6807+238G>C | 205 | 39 |

### Summary

Genotype of *XRCC1* 580C>T was not associated with RFS and OS in patients with cervical cancer who underwent radical surgery followed by adjuvant CRT. Similarly, none of genotype of *XRCC1* 1196A>G, *ATM* 3078-77C>T, *ATM* 5557G>A and *ATM* 6807+238G>C was associated with RFS or OS. In addition, none of haplotype of SNPs was associated with RFS or OS. None of genotype of SNPs was associated with completion of CRT. Low concordance rate between cancer and germline genotype was identified in cervical cancer. Age, stage, histologic type, LVSI and parametrial invasion were independently associated with RFS. Clinical tumor size, resection margin, lymph node involvement and completion of CRT were associated with OS in patients who underwent radical surgery followed by adjuvant CRT.

## Data Availability

Data can be provided upon reasonable requests.

## Acknowledgments

This work was supported by the intramural research fund of Korea Institute of Radiological & Medical Sciences.

## Notes

### Competing Interest Statement

The authors have declared no competing interest.

### Author Declarations

The KIRAMS Institutional Review Board approved the study protocol and waived the acquisition of informed consent(K-1101-002-058).

## References

1. Kamangar F, Dores GM, Anderson WF. Patterns of cancer incidence, mortality, and prevalence across five continents: defining priorities to reduce cancer disparities in different geographic regions of the world. J Clin Oncol 2006;24:2137–50.

2. Peters WA, 3rd, Liu PY, Barrett RJ, 2nd, et al. Concurrent chemotherapy and pelvic radiation therapy compared with pelvic radiation therapy alone as adjuvant therapy after radical surgery in high-risk early-stage cancer of the cervix. J Clin Oncol 2000;18:1606–13.

3. Kim K, Kang SB, Chung HH, Kim JW, Park NH, Song YS. Comparison of chemoradiation with radiation as postoperative adjuvant therapy in cervical cancer patients with intermediate-risk factors. Eur J Surg Oncol 2009;35:192–6.

4. Song S, Song C, Kim HJ, et al. 20year experience of postoperative radiotherapy in IB-IIA cervical cancer patients with intermediate risk factors: Impact of treatment period and concurrent chemotherapy. Gynecol Oncol 2012;124:63–7.

5. Sedlis A, Bundy BN, Rotman MZ, Lentz SS, Muderspach LI, Zaino RJ. A randomized trial of pelvic radiation therapy versus no further therapy in selected patients with stage IB carcinoma of the cervix after radical hysterectomy and pelvic lymphadenectomy: A Gynecologic Oncology Group Study. Gynecol Oncol 1999;73:177–83.

6. Alsner J, Andreassen CN, Overgaard J. Genetic markers for prediction of normal tissue toxicity after radiotherapy. Semin Radiat Oncol 2008;18:126–35.

7. Parliament MB, Murray D. Single nucleotide polymorphisms of DNA repair genes as predictors of radioresponse. Semin Radiat Oncol 2010;20:232–40.

8. Kim K, Kang SB, Chung HH, Kim JW, Park NH, Song YS. XRCC1 Arginine194Tryptophan and GGH-401Cytosine/Thymine polymorphisms are associated with response to platinum-based neoadjuvant chemotherapy in cervical cancer. Gynecol Oncol 2008;111:509–15.

9. Carles J, Monzo M, Amat M, et al. Single-nucleotide polymorphisms in base excision repair, nucleotide excision repair, and double strand break genes as markers for response to radiotherapy in patients with Stage I to II head-and-neck cancer. Int J Radiat Oncol Biol Phys 2006;66:1022–30.

10. Warnecke-Eberz U, Vallbohmer D, Alakus H, et al. ERCC1 and XRCC1 gene polymorphisms predict response to neoadjuvant radiochemotherapy in esophageal cancer. J Gastrointest Surg 2009;13:1411–21.

11. Sakano S, Wada T, Matsumoto H, et al. Single nucleotide polymorphisms in DNA repair genes might be prognostic factors in muscle-invasive bladder cancer patients treated with chemoradiotherapy. Br J Cancer 2006;95:561–70.

12. Tu HF, Chen HW, Kao SY, Lin SC, Liu CJ, Chang KW. MDM2 SNP 309 and p53 codon 72 polymorphisms are associated with the outcome of oral carcinoma patients receiving postoperative irradiation. Radiother Oncol 2008;87:243–52.

13. Yoon SM, Hong YC, Park HJ, et al. The polymorphism and haplotypes of XRCC1 and survival of non-small-cell lung cancer after radiotherapy. Int J Radiat Oncol Biol Phys 2005;63:885–91.

14. Angele S, Romestaing P, Moullan N, et al. ATM haplotypes and cellular response to DNA damage: association with breast cancer risk and clinical radiosensitivity. Cancer Res 2003;63:8717–25.

15. Goetz MP, Rae JM, Suman VJ, et al. Pharmacogenetics of tamoxifen biotransformation is associated with clinical outcomes of efficacy and hot flashes. J Clin Oncol 2005;23:9312–8.

16. Marsh S, Mallon MA, Goodfellow P, McLeod HL. Concordance of pharmacogenetic markers in germline and colorectal tumor DNA. Pharmacogenomics 2005;6:873–7.

17. Mort R, Mo L, McEwan C, Melton DW. Lack of involvement of nucleotide excision repair gene polymorphisms in colorectal cancer. Br J Cancer 2003;89:333–7.

18. Rae JM, Cordero KE, Scheys JO, Lippman ME, Flockhart DA, Johnson MD. Genotyping for polymorphic drug metabolizing enzymes from paraffin-embedded and immunohistochemically stained tumor samples. Pharmacogenetics 2003;13:501–7.

19. Schneider BP, Skaar TC, Sledge GW, Badve S, Li L, Flockhart DA. Analysis of angiogenesis genes from paraffin-embedded breast tumor and lymph nodes. Breast Cancer Res Treat 2006;96:209–15.

20. Liang CW, Lee YS, Marino-Enriquez A, Tsui K, Huang SH. The utility and limitation of single nucleotide polymorphism analysis on whole genome amplified mesenchymal tumour DNA in formalin fixed tumour samples. Pathology 2012;44:33–41.

